# Caregiver-mediated interventions to support self-regulation among infants and young children (0-5): A protocol for a realist review

**DOI:** 10.1101/2020.10.12.20211300

**Authors:** Amy Louise Finlay-Jones, Elaine Bennett, Jenny Downs, Sally Kendall, Keerthi Kottampally, Yi Huey Lim, Vincent Mancini, Rhonda Marriott, Helen Milroy, Monique Robinson, Justin Dean Smith, Lauren Wakschlag, Jeneva L. Ohan

**Affiliations:** Telethon Kids Institute, Nedlands, Australia; School of Psychology, Curtin University, Bentley, Australia; School of Medicine, The University of Western Australia, Crawley, Australia; Ngala, Kensington, Australia; School of Nursing and Midwifery, University of Notre Dame, Fremantle, Australia; School of Physiotherapy, Curtin University, Bentley, Australia; Centre for Health Services Studies, University of Kent, Canterbury, United Kingdom; Ngangk Yira Aboriginal Research Centre, Murdoch University, Murdoch, Australia; School of Psychological Sciences, University of Western Australia, Crawley, Australia; School of Medicine, University of Utah, Utah, United States of America; Feinberg School of Medicine, Northwestern University, United States of America

## Abstract

**Background and Objectives:** Self-regulation is a modifiable protective factor for lifespan mental and physical health outcomes. Early caregiver-mediated interventions to promote infant and child regulatory outcomes prevent long-term developmental, emotional, and behavioural difficulties and improve outcomes such as school readiness, educational achievement, and economic success. To harness the population health promise of these programmes, there is a need for more nuanced understanding of the impact of these interventions. The aim of this realist review is to understand how, why, under which circumstances, and for whom, early caregiver-mediated interventions improve infant and child self-regulation. The specific research questions guiding this review were based on consultation with families and community organizations that provide early childhood and family services.

**Design, Methods and Analysis:** Realist reviews take a theory-driven and iterative approach to evidence synthesis, structured around continuous refinement of a programme theory. Programme theories specify context-mechanism-outcome configurations to explain what works, for whom, under which circumstances, and how. Our initial programme theory is based on prior work in this field and will be refined through searching peer-reviewed and grey literature to identify relevant evidence. A working group, comprising service users, community organization representatives, representatives from specific populations, clinicians, and review team members will be formed to guide the evidence synthesis and interpretation, as well as the development and dissemination of recommendations based on the findings of the review. The review will involve searching: (1) electronic databases (e.g. EMBASE, Medline, PsycInfo), (2) connected papers, articles and citations, and (3) grey literature. Decisions to include evidence will be guided by judgements about their contribution to the programme theory and will be made by the research team, with input from the working group as required. Evidence synthesis will be reported using the RAMESES guidelines and disseminated through peer-reviewed publication.

**Trial registration number:** The protocol is registered with Open Science Framework https://osf.io/5ce2z/registrations

**Strengths and Limitations:**

- Uses a realist approach to provide insight into *what works, for whom, under which circumstances, and how* for caregiver-mediated interventions designed to promote self-regulation outcomes among infants and young children.
- Research questions were developed in consultation with families and service providers.
- Decision-making will be transparently documented, and all review materials made available on the Open Science Framework repository.
- The programme theory will be largely derived from peer-reviewed journal articles, and therefore may be subject to publication bias.

## Background

Self-regulation is an umbrella term used to refer to a variety of top-down and bottom-up processes implicated in the capacity to flexibly regulate behaviour, cognition, and emotion.^1 2^ Across developmental stages, healthy self-regulation is consistently associated with better mental health and adaptive functioning and improved human capital^3-6^ while self-regulatory difficulties are a risk factor for later mental and physical health problems.^5-8^ For example, self-regulatory difficulties in early childhood are associated with rapid weight gain through to early adolescence,^9^ while longitudinal studies have demonstrated that adults with no history of psychiatric diagnoses have significantly greater self-regulation in childhood than those with lifetime history of mental health problems.^4^ Differences in early self-regulation have been implicated in resilience to early adversity^10-12^ even when controlling for IQ and socioeconomic status.^12^ Self-regulation also predicts school readiness,^13^ academic achievement,^14^ self-worth, substance use, economic outcomes and criminal involvement.^6 15 16^

Interactions between biological and environmental factors shape regulatory outcomes over the course of infancy and early childhood.^17 18^ Among typically developing children, basic self-regulatory capacities begin to emerge in the first months of life^19 20^ and increase in complexity across development, demonstrating a developmental cascade effect that parallels changes in neurobiology.^21-23^ This dynamic and self-reinforcing process supports increasingly sophisticated skills for adaptive and executive functioning:^24 25^ as children develop greater self-regulatory skills, they are more able to effectively engage with their environment, thereby fostering more adaptive outcomes.^26 27^ It is important to note, however, that these adaptive developmental cascades assume that children develop in the context of manageable demands on their self-regulatory capacities. In the context of early adversity such as adverse prenatal exposures,^28 29^ socioeconomic stress, psychosocial threat, and neglect, cascade effects of adversity can work in the opposite way, undermining self-regulation and driving negative sequelae well into adulthood.^5 30^

### Promoting self-regulation through early intervention: the role of caregivers

Caregivers are instrumental in the development and maintenance of self-regulatory capacities among infants and young children.^31^ As they develop, infants transition from dependence on caregivers for co-regulation to more autonomous self-regulation via multiple, co-occurring socialization processes.^32^ Regardless of developmental stage, in situations where regulatory demands are overwhelming, co-regulation via caregivers is important to support infants and children to respond adaptively to their environment. However, much like other environmental influences, the impact of caregiver behaviour on child self-regulatory outcomes can be positive or negative: reviews and meta-analyses have found that the use of positive caregiving strategies (such as guiding and encouraging) is associated with more adaptive child self-regulation, while negative caregiving strategies (such as criticism) are associated with poorer self-regulatory outcomes.^31-34^ Furthermore, evidence that caregiving mediates the relationship between early adversity and regulatory outcomes in children^35 36^ suggests that supporting positive caregiving practices is particularly important for families experiencing adversity.

Evidence supports the assertion that caregiver-mediated interventions designed to foster self-regulation among infants and young children have potential to interrupt negative developmental cascades, and promote positive ones.^22^ Prior work by Murray and colleagues^37^ reviewing studies of self-regulation programmes between 1989-2013 demonstrated the breadth of this literature, although they found substantially fewer programmes targeting the birth to age 2 year group (k = 27) than those targeting 3-4 year olds (k = 75). Programmes that have been found to promote self-regulation in infants and young children include, but are not limited to, Attachment and Biobehavioural Catch-Up,^38 39^ Triple P,^40^ the Incredible Years,^41-43^ the Family Check-Up, ^44 45^ Parent-Child Interaction Therapy,^46-49^ Head Start,^50-52^ the Chicago School Readiness Project, ^53^ the Kids in Transition to School Project,^54^ and Tools of the Mind.^25 55-58^ However, inconsistencies in these programmes and their evaluation undermine capacity to draw inferences about how early self-regulation programmes work. For example, not all these programmes specifically target self-regulatory outcomes, there are differences used in the terminology used to describe the programme targets and components, and self-regulation outcomes are measured inconsistently across trials.^2 59^ Accordingly, it is unclear which components are considered key to promoting self-regulation outcomes and which are extraneous to this goal.^2^

### Factors influencing the effectiveness of caregiving programs to improve child self-regulation

Conceptual and methodological issues notwithstanding, these interventions have shown evidence of benefit for a range of parent and child outcomes across a variety of populations and settings.^60-63^ For example, there is evidence that early intervention can improve self-regulation among infants and children exposed to early risk factors, such as socioeconomic adversity,^64^ foster care,^38^ neglect,^39^ and pre-term birth.^65^ Early caregiving programmes may improve self-regulation outcomes when delivered remotely^66^ and in primary care settings. Together, these findings indicate a robustness of intervention effects across contexts. However, prior reviews and meta-analyses have also revealed important findings about the implementation of these interventions, including barriers to engagement and the circumstances under which such interventions do not work. For example, service location, incompatibility of programme delivery with work schedules, transport barriers, perceived stigma, and cost of delivery create considerable barriers to programme engagement and implementation.^67 68^ Synthesizing these data on practical considerations that influence engagement and outcomes is important to inform implementation strategies.

Despite the apparent robustness of self-regulation interventions across contexts, previous work has also highlighted how intervention characteristics interact with contextual factors to influence intervention effects. For instance, Rayno and McGrath ^69^ found that socioeconomic status and maternal mental health significantly predicted response to parenting interventions for child externalizing behaviour problems. Based on work demonstrating that socioeconomic stress adversely impacts caregiving behaviour by increasing caregiver distress, Rayno and McGrath suggested that one way of optimising intervention-context fit is to include additional components focusing on caregiver mental health when programmes are delivered to families with higher socioeconomic stress. Similarly, Harris, et al. ^66^ found that contact with an interventionist was necessary for technology-assisted caregiving interventions to effectively improve parent wellbeing in socioeconomically disadvantaged families. These findings illustrate the complexities of delivering ‘evidence-based’ caregiver-mediated self-regulation interventions in different settings for different target groups. The lack of synthesised understanding of how to respond to these complexities when delivering interventions can undermine translation. In reviewing the literature on caregiving interventions in paediatric primary care, Smith, et al. ^70^ found that the dearth of information regarding implementation methods and contexts undermined the potential to deliver scalable and equitable caregiving intervention models.

### Aims and Objectives

The aim of the current review is to extend existing evidence syntheses to provide insight into context-mechanism-outcome combinations underlying self-regulation interventions for 0-5 - year-old children. We aim to highlight *how* and *why* these programmes work, as well as to identify *for whom* and *under which circumstances* these programmes lead to positive outcomes for children and families.

Our objectives are as follows:

1. To synthesize insights from peer-reviewed and grey literature, stakeholder perspectives, and expert guidance regarding what works, for whom, under which circumstances, and how, for caregiver-mediated self-regulation interventions for infants and young children.
2. To develop a set of programme theories and corresponding evidence maps documenting relationships between intervention components, contextual factors, and mechanisms influencing outcomes of these interventions.
3. To produce guidelines for intervention development and implementation based on the evidence synthesis.

## Methods

### Realist Review

This review will use a realist approach to address questions around intervention mechanisms and implementation contexts and to generate insights into the question of what works, for whom, how, and in which settings. Realist synthesis is particularly useful where research is heterogenous, as is the case for the literature on early self-regulation interventions.^71^ The realist approach aims to generate policy-relevant findings for the purposes of decision-making in programme financing and implementation. Realist approaches are appropriate for complex interventions where intervention effects are context-dependent. While randomised controlled trials and meta-analyses help to answer the question of whether self-regulation interventions are effective or not, realist synthesis is a theory-driven approach that seek to determine why and how interventions do/do not work for different people in across different contexts.^72 73^

### Patient and Public Involvement

The need for this review emerged from a series of consultations with families with a child with developmental, emotional and/or behavioural difficulties, for the purposes of understanding their needs regarding early screening and support. These consultations highlighted that parental awareness of children’s developmental difficulties often preceded formal recognition of these difficulties by health professionals, and that due to the highly variable nature of early development, families were frequently told to wait and see whether more pronounced and stable child difficulties would emerge. Families described this as a time of high concern and stress, with caregiver mental health and family functioning undermined by the burden of child developmental, emotional and/or behavioural difficulties coupled with the anxiety of not knowing. Accordingly, a potential solution was to investigate early intervention approaches that were *cross-syndrome* or *transdiagnostic*; that is, they target risk and/or protective factors that are implicated in a wide range of developmental, emotional and behavioural difficulties, and can be implemented prior to formal diagnosis.

Self-regulation is one such transdiagnostic factor that is meaningful to caregivers and service providers, given that child self-regulatory difficulties are a common reason that parents seek support from health professionals and family services. Our community partners (organisations providing family and early childhood services in Australia) identified a gap in the availability of evidence-based self-regulation interventions for infants and toddlers and expressed a desire to understand more about which interventions should be recommended for which families under which circumstances. Of particular interest to our community partners is the appropriateness of different self-regulation interventions for Aboriginal and Torres Strait Islander (Indigenous) families, Culturally and Linguistically Diverse (CALD) families, families experiencing socioeconomic disadvantage, and families living in rural and remote areas.

Together, the input from families and community partners directly shaped the focus of the proposed review. The need to synthesise insights into *what works, for whom, under which circumstances and how* for early self-regulation interventions is guided by our community partners’ interest in optimising intervention strategies for the multiple communities they serve. It is also of importance to guide families, who, in the absence of formal recognition and guidance regarding child difficulties, are often required to make decisions about which types of strategies and services are most appropriate for their needs.

We have planned for ongoing involvement of families and community partners in the review process. We will establish a working group with a balance of caregivers and representatives from community organisations to provide input into all stages of the review. The group will comprise representatives from community organizations (early childhood and family services), practitioners, service users (i.e. parents with young children), research and clinical experts in the field of infant and child self-regulation, as well as members of the review team. The working group will play a key role in interpreting the findings of the review, developing theory, and establishing consensus. They will also play a key role in developing the recommendations and guiding translation of the findings into policy and practice. We anticipate that the findings of the review will be used to guide intervention development and/or implementation as part of our ongoing work in this field. Families and community partners would also be closely involved as co-design partners in any intervention development or the generation of implementation recommendations arising from this review.

## Research Questions

We will use a realist synthesis approach to answer the following questions:

1. What are the key contextual factors (for example, implementation strategies, setting, mode of delivery, and population characteristics) which influence the success or failure of early caregiving interventions to support self-regulation?
2. What are the core intervention components and key intervention mechanisms which, in the right contexts, lead to the success of early caregiving interventions to support self-regulation?
3. How do core components and key intervention mechanisms vary across age groups from 0-5 years?
4. What is known about what works to improve child self-regulatory outcomes among (i) Indigenous and CALD families; (ii) families with socioeconomic disadvantage; and (iii) families living in rural and remote areas? Specifically, how do contextual factors, core components, key mechanisms and outcomes vary across populations?
5. How might these findings influence future research, policy, and practice?

### Design

We will follow the five steps outlined by Pawson, et al. ^74^: (1) clarifying the review’s scope; (2) defining the search strategy; (3) selecting studies for inclusion in the review; (4) extracting data from included studies; and (5) evidence synthesis and recommendations. Steps 1-2 and 5 will be done in active consultation with working groups formed with families and other stakeholders. The review will be reported according to the Realist and MEta-narrative Evidence Synthesis: Evolving Standards (RAMESES) standards for realist syntheses.^75^ The review was registered with the Open Science Framework on the 10^th^ October, 2020. Project documents, including refinements to the search strategy, decisions regarding evidence inclusion and exclusion, and amendments to the protocol will be documented on the project site (https://bit.ly/34J7oY2).

The primary output of this review will be an evidence-based programme theory for early caregiving interventions to support infant and child self-regulation, highlighting the key mechanisms of these interventions and the contextual factors that influence their effectiveness across populations and settings. We aim to use the findings of this review to generate a set of recommendations for the development and delivery of early caregiving programmes to optimise intervention effectiveness, fidelity, and equity. Recommendations will concern core components, implementation principles and strategies, and considerations regarding setting and mode of delivery. We also aim to use the findings of this review to inform local intervention development and implementation efforts.

### Working Definitions and Preliminary Scope

For the purposes of this review, ‘early self-regulation interventions’ are those that are delivered from the time children are born up to and including 5 years of age. We will focus on caregiver-mediated interventions given that most early self-regulation interventions for this age group focus on caregivers^76^ and because this focus aligns with the priorities and interests of the families and community organisations we consulted with. Following Murray and colleagues,^37^ we will include studies of caregiving interventions that either explicitly target infant/child self-regulation or those that measure infant or child self-regulation as an outcome. As interventions that promote self-regulation have been referred to in the literature using various terms, we will also focus on interventions that aim to promote children’s executive functioning and emotion regulation skills and reduce challenging behaviour. We will include universal or targeted interventions delivered across all contexts and settings.

### Step 1: Clarifying the scope of the review and developing a programme theory

Realist reviews are theory-driven, using an iterative approach to selecting, developing, and refining programme theories to explain why and how interventions work for different people in different contexts.^72 74^ Thus, the first stage of the review will involve clarifying the review scope and defining the initial programme theory. Given the inconsistencies in terminology used across self-regulation interventions, an initial step is to identify key constructs associated with self-regulation intervention and develop a list of exemplar programs. These will be identified through initial scoping of the literature using a broad set of search terms (see Table 1 for an example of the Medline search strategy). In addition to searching peer reviewed and grey literature, we will liaise with the working group to identify other forms of evidence that do not take the form of peer reviewed studies, such as policy documents. At the scoping stage, we will focus on programmes that have been developed for infants and young children, 0-5 years, that involve caregiver-mediated strategies to promote child self-regulation and/or that measure infant and child self-regulation as an outcome. As the focus of the realist review is on theory development, we will include intervention studies with and without a comparator condition.

**Table 1.**
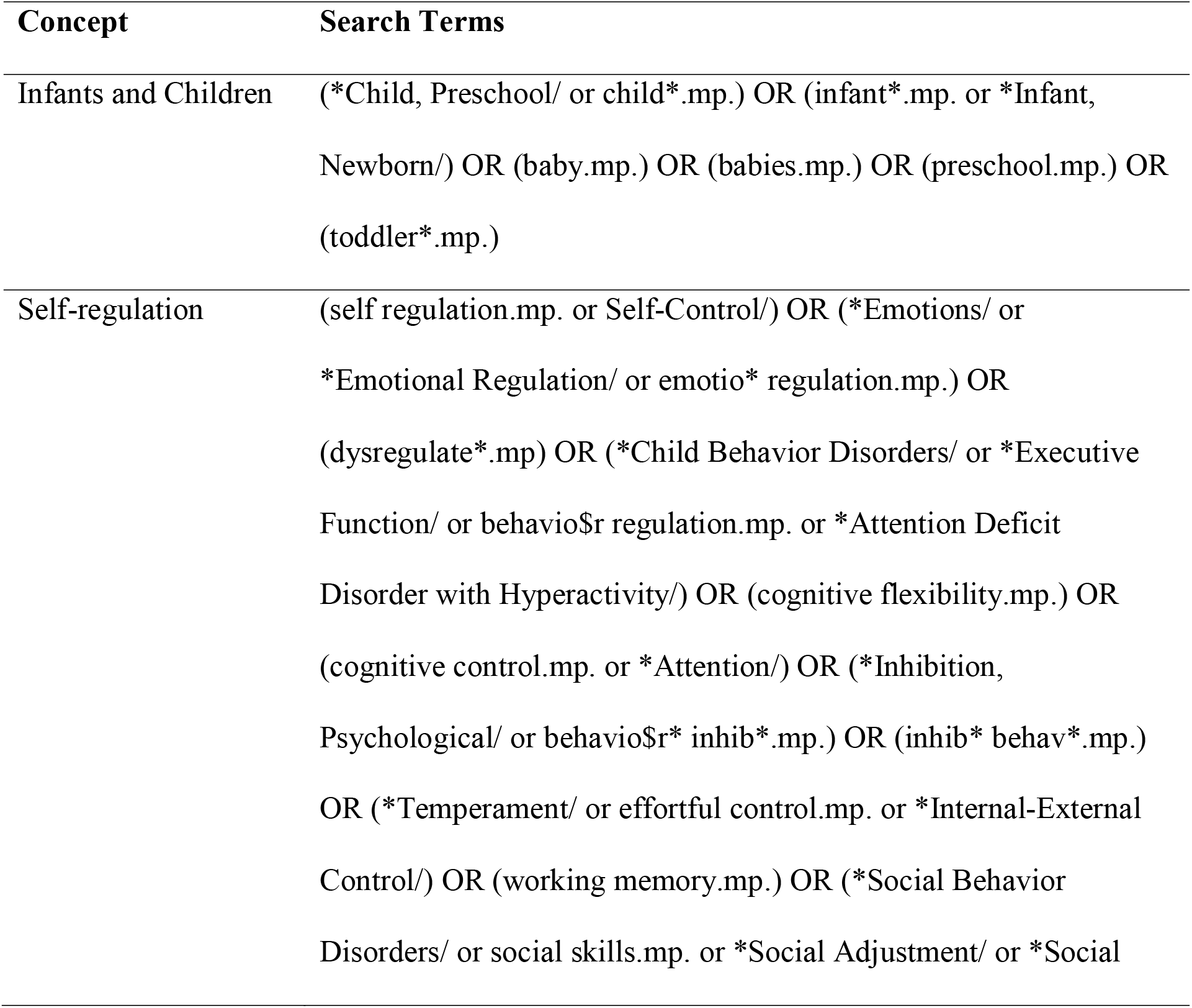
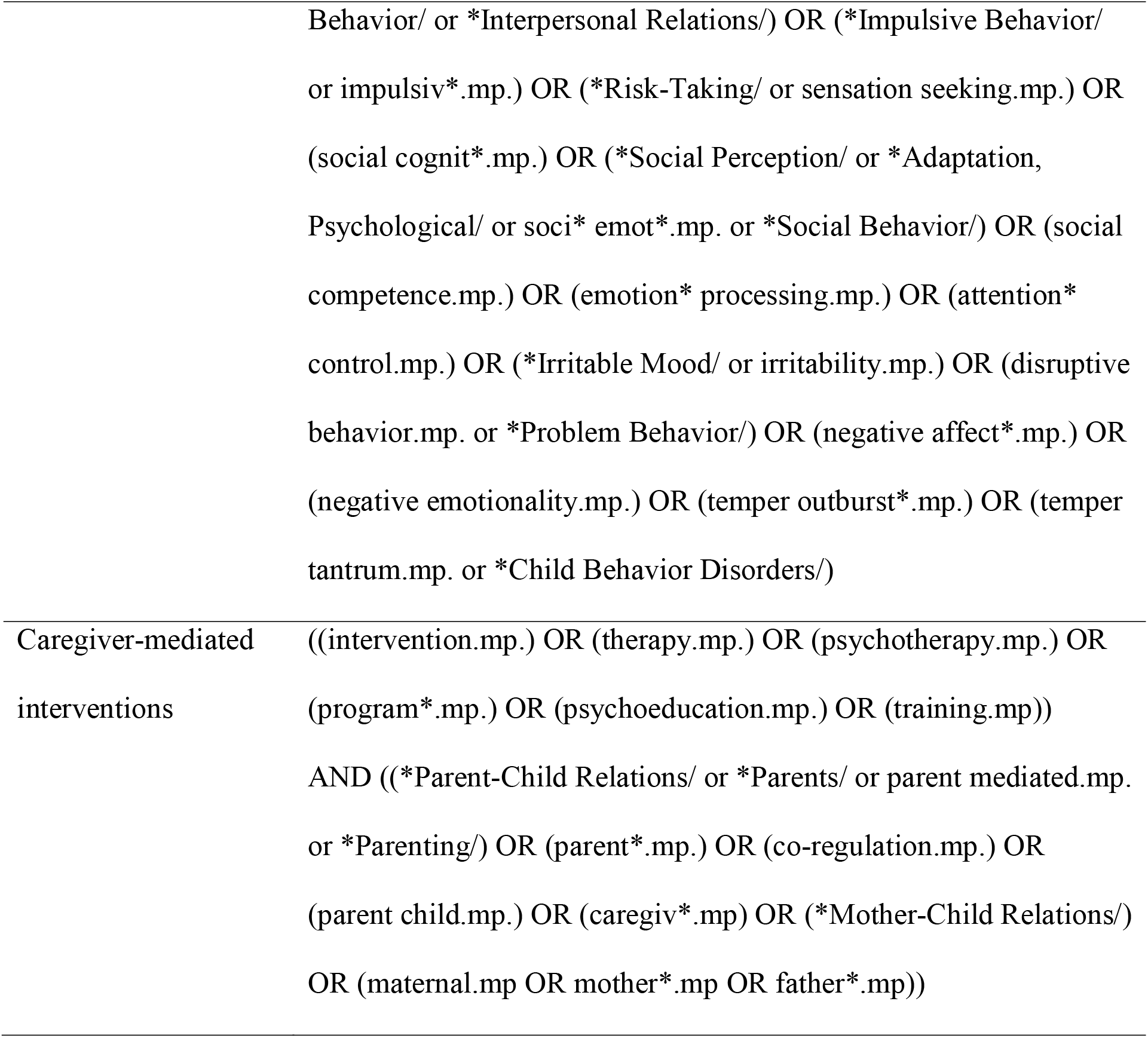
Medline search strategy for scoping stage.

| Concept | Search Terms |
| --- | --- |
| Infants and Children | (*Child, Preschool/ or child*.mp.) OR (infant*.mp. or *Infant, Newborn/) OR (baby.mp.) OR (babies.mp.) OR (preschool.mp.) OR (toddler*.mp.) |
| Self-regulation | (self regulation.mp. or Self-Control/) OR (*Emotions/ or *Emotional Regulation/ or emotio* regulation.mp.) OR (dysregulate*.mp) OR (*Child Behavior Disorders/ or *Executive Function/ or behavio\$r regulation.mp. or *Attention Deficit Disorder with Hyperactivity/) OR (cognitive flexibility.mp.) OR (cognitive control.mp. or *Attention/) OR (*Inhibition, Psychological/ or behavio\$r* inhib*.mp.) OR (inhib* behav*.mp.) OR (*Temperament/ or effortful control.mp. or *Internal-External Control/) OR (working memory.mp.) OR (*Social Behavior Disorders/ or social skills.mp. or *Social Adjustment/ or *Social |
|  | Behavior/ or *Interpersonal Relations/) OR (*Impulsive Behavior/ or impulsiv*.mp.) OR (*Risk-Taking/ or sensation seeking.mp.) OR (social cognit*.mp.) OR (*Social Perception/ or *Adaptation, Psychological/ or soci* emot*.mp. or *Social Behavior/) OR (social competence.mp.) OR (emotion* processing.mp.) OR (attention* control.mp.) OR (*Irritable Mood/ or irritability.mp.) OR (disruptive behavior.mp. or *Problem Behavior/) OR (negative affect*.mp.) OR (negative emotionality.mp.) OR (temper outburst*.mp.) OR (temper tantrum.mp. or *Child Behavior Disorders/) |
| Caregiver-mediated interventions | ((intervention.mp.) OR (therapy.mp.) OR (psychotherapy.mp.) OR (program*.mp.) OR (psychoeducation.mp.) OR (training.mp))<br>AND ((*Parent-Child Relations/ or *Parents/ or parent mediated.mp. or *Parenting/) OR (parent*.mp.) OR (co-regulation.mp.) OR (parent child.mp.) OR (caregiv*.mp) OR (*Mother-Child Relations/) OR (maternal.mp OR mother*.mp OR father*.mp)) |

In the realist approach, programme theories are comprised of ‘context’ (C), ‘mechanism’ (M) and ‘outcome’ (O) – referred to here as the C-M-O framework. Interventions are thought to impact outcomes by altering context (e.g., increasing caregiver knowledge and skills), thereby influencing the mechanisms (e.g., increasing responsive caregiving behaviour) that drive outcomes. Prior reviews have demonstrated that caregiving programs to improve child self-regulation include a vast array of program components. For example, effective interventions for preschool-age children have included components to increase proactive parenting, parent involvement, positive behaviour management, parental guidance, and limit-setting.^45 51^ It is unclear whether these different components influence self-regulatory outcomes via a smaller subset of shared mechanisms. For example, Sandler, et al. ^77^ suggest that three main factors account for long-term effects of parenting programmes: parenting skills, self-efficacy, and parent mental health.

In the scoping stage, we aim to identify existing programme theories, or components of programme theories, from the literature and other evidence documents. Following Coles, et al. ^78^ programme theories may be identified from literature describing the hypothesized causal mechanisms of the programs, their theoretical bases, and descriptions of the relationship between programme activities and outcomes. These theories will be synthesized to generate a programme theory structured around the C-M-O framework. We will also consult with the working group to identify experts and key stakeholders to input to the programme theory. The resulting theoretical, explanatory model and its component contexts, mechanisms and outcomes of interest will be used as the framework for the succeeding stages of the review.

### Step 2. Refining the search strategy

The refined search strategy will be based on the emerging programme theory. Given the vast array of potential contexts, mechanisms, and outcomes that may emerge from the self-regulation intervention literature, we will consult with the working group to decide which C-M-O variables should be the primary focus of the review. Once this has been agreed, we will systematically search for literature to extend and refine the scope of studies identified in Step 1, with the aim of testing and refining the programme theory. This approach aligns with the iterative nature of the realist methodology.^74^ Electronic database searching in Ovid Embase, PsycInfo, Medline and Web of Science^79^ will be carried out using keywords based on the interventions, concepts, mechanisms, theories, and outcomes identified in the scoping stage. We will identify additional studies for inclusion by handsearching reference lists of included papers to find connected texts and by searching the grey literature. The search will be a multi-stage process that integrates consultation with the working group and other key stakeholders. A series of Endnote libraries will be created to document the results of each stage of the search process.

### Step 3: Selecting data sources for inclusion in the review

Selection of data sources for inclusion in the review will primarily be based on their relevance to the C-M-O components of the programme theory. Several different types of evidence can be integrated within the scope of a realist review^75^; accordingly, we anticipate that data sources will include peer-reviewed journal articles, policy documents, and programme manuals. Two research team members will independently screen titles and abstracts of the data sources against the inclusion and exclusion criteria (or refined versions of these criteria developed following the initial search). Data sources will be independently ranked by two members of the team, according to their relevance to their programme theory and the rigour of the findings. The rigour dimension will be appraised by the review team members based on how robustly the methods of a given data source (including entire studies and components within a study) support the conclusions drawn from it. The review team will use these rankings to guide selection of data sources, in consultation with the working group. Excel spreadsheets will be used to track decisions and rationale for the inclusion and exclusion of specific studies.

### Step 4: Extracting data from included sources

According to the realist approach, extracted data should be used to determine whether programme theories and their components are meaningful and productive.^80^ Data sources and extracted data will therefore be included based on their capacity to test and refine the emerging programme theory. Extracted data will be mapped against the programme theory and research questions, with the overarching aim of identifying the core components of effective self-regulation interventions across contexts. We will design data extraction forms based on the C-M-O framework and using the interventions, contexts, mechanisms, theories, and outcomes identified in the scoping stage. It is anticipated that, at a minimum, extracted data will include study authors, year of publication, study country; study design; sample characteristics and inclusion criteria; intervention components, mode of delivery, and characteristics of intervention facilitators, as well as data on the context in which the intervention is studied, and predictors, mediators, and moderators of intervention outcome. Finally, we will extract data on outcomes: our primary outcome of interest is infant and child self-regulation, however we are also interested in related outcomes such as infant and child mental health, family functioning, caregiver mental health, self-regulation, caregiving behaviours, and self-efficacy. Quality appraisal (separate from evaluations of relevance and rigour) will be undertaken to summarise the overall quality of the included studies. We will use the Cochrane RoB 2 revised risk-of-bias tool^81^ for randomized trials, the Cochrane ROBINS-I tool^82^ for non-randomized intervention studies, and the 32-item Consolidated Criteria for Reporting Qualitative Research.^83^

### Step 5: Evidence synthesis and recommendations

The focus of evidence synthesis will be on the testing and refinement of the programme theory. In this stage of the review, we will code interventions using the C-M-O framework and generate a series of C-M-O configurations that can be applied to different populations across different settings. We will undertake qualitative data synthesis using NVivo, and map interventions against C-M-O variables using an evidence mapping spreadsheet developed for the purposes of this review. Where relevant, summary statistics will be used to characterise the included studies. We will use a narrative approach to synthesise findings and report these according to the RAMESES guidelines.

### Generation of Recommendations and Dissemination of Findings

A draft summary of the findings and recommendations will be reviewed by the working group and key stakeholders, who will provide input into their relevance and meaning for policy and practice applications. A final list of recommendations for the development and implementation of caregiver-mediated interventions to support self-regulation among infants and young children will be determined by consensus of the working group. Findings and recommendations will be disseminated through a report and policy brief, journal articles, and stakeholder presentations. We will also work with the caregivers in our working group and broader community networks to develop evidence summaries that are useful to families.

## Discussion

### Importance of the Research

Self-regulation is a cornerstone of healthy development,^5^ and early interventions to promote self-regulation have potential to support adaptive outcomes across the lifespan.^37^ This realist review will provide important insights into how caregiver-mediated interventions for infants and young children promote self-regulation outcomes and improve long-term functioning. Understanding how such interventions work, who they work for, and under which contexts, can help to optimize intervention and implementation strategies. Documenting how well the existing evidence supports the programme theory will also aid in refining theoretical understandings of caregiver-mediated interventions for infants and young children and highlight key gaps in the research. In addition, identifying mechanisms underlying early caregiver-mediated programmes to improve self-regulation process, will enable us to design experimental studies to test these mechanisms in the future.

We anticipate that the findings of this review will provide key practical advice for health professionals and community service providers working to provide early childhood and family services, as well as service users, policymakers, and researchers. Our intention is to provide a trans-professional explanation of how, why, and under which circumstances caregiver-led self-regulation interventions can best support children’s development. We will work with the range of stakeholders in our working group to determine the best way to summarise and disseminate the findings from the review so it can be used to guide practical decisions around implementation.

## Supporting information

PRISMA-P Checklist

## Data Availability

Review documents will be made publicly available on OSF

https://osf.io/5ce2z/?view_only=d9a4b4d5487c403f8ff2396d2f11814a

## Contributors

AFJ conceptualised the study and led the design and drafting of the review protocol and manuscript. KK, JO and LW helped to draft the search protocol. All authors provided feedback on the review protocol and provided comments to improve the manuscript. All authors have read and approved the final manuscript. AFJ acts as guarantor for the review.

