## Supplementary material for "Caregiver-mediated interventions to support self-regulation among infants and young children (0-5): A protocol for a realist review": PRISMA-P Checklist

| Section and topic | Item No | Checklist item | | Page |
| --- | --- | --- | --- | --- |
| **Administrative information** |  | |  |  |
| Title: |  |  | |  |
| Identification | 1a | Identify the report as a protocol of a realist review | | 1 |
| Update | 1b | If the protocol is for an update of a previous systematic review, identify as such | | N/A |
| Registration | 2 | If registered, provide the name of the registry (such as PROSPERO) and registration number | | 4 |
| Authors: |  |  | |  |
| Contact | 3a | Provide name, institutional affiliation, e-mail address of all protocol authors; provide physical mailing address of corresponding author | | 1-2 |
| Contributions | 3b | Describe contributions of protocol authors and identify the guarantor of the review | | 21 |
| Amendments | 4 | If the protocol represents an amendment of a previously completed or published protocol, identify as such and list changes; otherwise, state plan for documenting important protocol amendments | | 13 |
| Support: |  |  | |  |
| Sources | 5a | Indicate sources of financial or other support for the review | | 21 |
| Sponsor | 5b | Provide name for the review funder and/or sponsor | | N/A |
| Role of sponsor or funder | 5c | Describe roles of funder(s), sponsor(s), and/or institution(s), if any, in developing the protocol | | 21 |
| **Introduction** |  | |  |  |
| Rationale | 6 | Describe the rationale for the review in the context of what is already known | | 5-8 |
| Objectives | 7 | Provide an explicit statement of the question(s) the review will address with reference to participants, interventions, comparators, and outcomes (PICO) | | 9 |
| **Methods** |  | |  |  |
| Eligibility criteria | 8 | Specify the study characteristics (such as PICO, study design, setting, time frame) and report characteristics (such as years considered, language, publication status) to be used as criteria for eligibility for the review | | 14 |
| Information sources | 9 | Describe all intended information sources (such as electronic databases, contact with study authors, trial registers or other grey literature sources) with planned dates of coverage | | 17 |
| Search strategy | 10 | Present draft of search strategy to be used for at least one electronic database, including planned limits, such that it could be repeated | | 15-16 |
| Study records: |  |  | |  |
| Data management | 11a | Describe the mechanism(s) that will be used to manage records and data throughout the review | | 17-18 |
| Selection process | 11b | State the process that will be used for selecting studies (such as two independent reviewers) through each phase of the review (that is, screening, eligibility and inclusion in meta-analysis) | | 17-18 |
| Data collection process | 11c | Describe planned method of extracting data from reports (such as piloting forms, done independently, in duplicate), any processes for obtaining and confirming data from investigators | | 18 |
| Data items | 12 | List and define all variables for which data will be sought (such as PICO items, funding sources), any pre-planned data assumptions and simplifications | | 18 |
| Outcomes and prioritization | 13 | List and define all outcomes for which data will be sought, including prioritization of main and additional outcomes, with rationale | | 18-19 |
| Risk of bias in individual studies | 14 | Describe anticipated methods for assessing risk of bias of individual studies, including whether this will be done at the outcome or study level, or both; state how this information will be used in data synthesis | | 18-19 |
| Data synthesis | 15a | Describe criteria under which study data will be quantitatively synthesised | | 19 |
|  | 15b | If data are appropriate for quantitative synthesis, describe planned summary measures, methods of handling data and methods of combining data from studies, including any planned exploration of consistency (such as I^2^, Kendall’s τ) | | 19 |
|  | 15c | Describe any proposed additional analyses (such as sensitivity or subgroup analyses, meta-regression) | | N/A |
|  | 15d | If quantitative synthesis is not appropriate, describe the type of summary planned | | 19 |
| Meta-bias(es) | 16 | Specify any planned assessment of meta-bias(es) (such as publication bias across studies, selective reporting within studies) | | N/A |
| Confidence in cumulative evidence | 17 | Describe how the strength of the body of evidence will be assessed (such as GRADE) | | N/A |
